# Effect of Brief Dog-Assisted Interventions on Stress Biomarkers: A Systematic Review

**DOI:** 10.1101/2023.12.10.23299796

**Authors:** Caroline Faucher, Anna Behler, Megan Campbell, Renate Thienel

**Affiliations:** School of Psychological Sciences, The University of Newcastle; School of Medicine and Public Health, The University of Newcastle

**Keywords:** Animal-assisted therapy, dog-assisted intervention, stress biomarkers

## Abstract

Despite the growing popularity of dog-assisted interventions (DAI), limited empirical evidence exists on the effect of this therapeutic approach on biological stress responses. This systematic review focused on single instances of a dog-assisted intervention to specifically assess treatment outcomes on stress biomarkers in participants across the lifespan. A total of 26 studies met the inclusion criteria. Cortisol level, blood pressure and heart rate were the most frequently analyzed biomarkers. Evidence to date partially supports the notion that a brief therapeutic intervention with the assistance of a dog may influence the autonomic stress response given that the majority of studies found significant changes in stress biomarkers in groups receiving DAI compared to control groups. However, the present review failed to reach a conclusion regarding the effect of DAI on stress biomarkers within a specific population or settings mostly due to the heterogeneity of the studies. More studies of high quality, and with more transparent and standardized protocols are necessary to further understand the physiological outcomes of DAI in order to develop targeted, evidence-based interventions.

## Effect of Dog-Assisted Interventions on Stress Biomarkers: A Systematic Review

It is well established that humans and dogs have a strong bond. Both seek out interaction from one another. Throughout history, dogs have supported humans in many ways and vice versa (Chambers et al., 2020). Dogs have accompanied soldiers on battlefields, worked alongside farmers, protected people from danger and kept them company at home. In return, humans have provided food, shelter, companionship and ensured their welfare. This mutually beneficial relationship between humans and animals is commonly referred to as the human-animal bond (Beck, 2014). Recently, this bond with dogs has been increasingly drawn upon to assist in reducing stress as part of the therapeutic process (Chandler, 2012; Machova et al., 2020). However, despite the growing popularity of dog-assisted interventions (DAI), limited empirical evidence exists on their effect on the biological stress response. A better understanding of the underlying therapeutic outcomes of DAI is necessary to develop targeted, evidence-based interventions.

### The Foundation of Dog-Assisted Interventions

The human-animal bond is widely thought to be rooted in evolutionary development (Beck, 2014). According to Beck, relationships with animals fulfil some core biological, psychological, and physiological needs. This is supported by three theories, which are frequently referred to in the literature on the human-animal bond. Firstly, the biophilia hypothesis suggests that humans have an innate tendency to seek connections with other forms of life (Wilson, 1986). Secondly, the social support theory emphasizes the importance of social relationships for the well-being and physical health of humans (Cullen & Wilcox, 2010). Finally, the theory of attachment (Bowlby, 1958) seeks to explain the deep and enduring emotional relationship between humans and their companion animals. Conversely, it is the dogs’ understanding of human behavior that predisposed them to interact with people (Horowitz, 2009).

The intrinsic benefits of pet ownership (Friedmann et al., 1980) is what boosted interest in the human-animal bond. For instance, one of the first pieces of biological evidence that supported this bond showed that when owners pet their dog, there was an increase in oxytocin in both the animal and the human (Odendaal, 2001). These findings set the stage for the inclusion of animals in various therapeutic and educational interventions (Fine & Beck, 2019).

### Dogs-Assisted Interventions to Reduce Stress

The International Association of Human-Animal Interaction Organizations (IAHAIO; 2013) defines animal-assisted interventions (AAI) as any goal-directed intervention that intentionally incorporates animals in health, educational, or social settings for a human’s therapeutic or educational benefit. Animal-assisted therapy is more specific and sits under the broader AAI category. It refers to goal-directed interventions that complement conventional treatment and are delivered by credentialed health or human service professionals (Fine, 2019).

Research in this field is relatively new despite the growing interest in AAI as a therapeutic modality and respectable form of complementary intervention. Still, several studies have yielded positive outcomes, particularly in the realm of stress reduction. The American Psychological Association (2020) defines stress as the physiological and psychological response to stressors. The Mental Health Foundation study (2018) conducted in the United Kingdom found that, within one year, nearly three quarters of adults had felt so stressed they were overwhelmed or unable to cope. In turn, stress has been associated with a range of mental health and physiological issues such as depression, anxiety, cardiovascular diseases, and diabetes (Mariotti, 2015).

AAIs targeting stress mostly involve dogs and are often referred to as canine or dog-assisted interventions (DAI is used throughout this review for simplicity). In a randomized controlled trial, for instance, regular visits from therapy dogs to pediatric oncology patients contributed to a reduction in state anxiety in children and decreased parental stress in the initial stages of cancer treatment (McCullough et al., 2018). Other studies have investigated the effects of a DAI on the well-being of students. For instance, an intervention as brief as 20 minutes with a dog trained to assist with the delivery of therapeutic interventions was found to alleviate stress and improve the mood of university students who were randomly assigned to the therapy condition (Grajfoner et al., 2017). While there are fewer studies on the effect of DAI on older adults, a randomized pilot study by Branson et al. (2020) suggested that brief interventions could reduce psychological stress in older intensive care patients.

While the evidence presented thus far supports the idea that DAI can reduce stress, the majority of studies have used self-report methods or behavioral observations, thus mainly reporting effects on psychological stress. In contrast, few studies have used objective biomarkers to measure the biological processes underlying the impact of AAI on the physiological stress response, to complement these subjective findings. In those that have, results have been mixed. For example, a meta-analysis on the effect of pet-assisted interventions on the physiological and subjective stress response, found a significant effect on heart rate, but not on blood pressure (Ein et al., 2018). The biomarkers in the analysis were limited to heart rate and blood pressure and did not include other commonly used stress biomarkers such as cortisol levels. Furthermore, the analysis was broad, and included many different types of animals and studies designs.

Evidence of the effect of dog-assisted therapy on the autonomic stress response remains poorly understood. Considering that one of the criticisms of the broader field is that AAI is tainted by exaggeration and misrepresentation (Fine et al., 2019), there is a great need for research looking at high-level scientific evidence to empirically assess specific animal-assisted interventions. As such, this review investigated the use of biomarkers in randomized controlled trials of brief stress interventions with the assistance of dogs. Having a better understanding of the physiological changes underlying the therapeutic outcomes of DAI across the lifespan will enable the design of better-targeted interventions.

## Methods

This systematic review is guided by the PRISMA 2020 Statement (Page et al., 2021) to ensure methodological consistency and transparency. The review was previously registered on PROSPERO (CRD42022313164). However, it was later decided that study designs would be expanded to not only include randomized controlled trials (RCT; see new criteria below).

### Eligibility Criteria

The search was limited to English-language articles published in peer-reviewed journals, (excluding dissertations), up until November 2023 in two databases commonly used in psychology: PsycINFO and PubMed. Studies were eligible if they were measuring the effects of a brief animal-assisted intervention as defined by IAHAIO (2013; see earlier definition). Interventions constituted of a single therapy instance with at least one group undergoing a control condition. Animals used in the interventions were limited to dogs. Group therapy was excluded. Only articles containing at least one objective biomarker measure commonly associated with stress were included, such as changes in hormone or protein levels, blood pressure, galvanic skin response, heart rate, electro-physiological signals, and neural activity (Bagheri & Power, 2020; Gordon & Mendes, 2021; Hek et al., 2013; Khan, 2020; Satti et al., 2021; Taelman et al., 2009). There were no restrictions on the population. In summary, the review focused on a single instance of a dog-assisted intervention to specifically assess treatment outcomes on stress biomarkers in participants across the lifespan.

### Search Strategy

A preliminary literature search was conducted to identify relevant keywords for the review. Of notable importance was the inclusion of a broad range of intervention terms due to the variability of terms used to describe DAI. The refined search for literature included in the review made use of Boolean operator ‘OR’ within each element, the Boolean operator ‘AND’ to link elements together and the asterisk to find words starting with the same letters. Table 1 summarizes the terms used.

**Table 1:** Terms Used to Search Literature.

| Search components | Search terms |
| --- | --- |
| Intervention | "animal-assisted" or "dog-assisted" or "canine-assisted" or "pet-assisted" or "therapy dog*" or "therapy pet" or "therapy animal*" or "animal assisted" |
| Outcome | imaging or encephalogra* or eeg or saliva or "heart rate" or cortisol or "blood pressure" or galvanic or "neural activity" or autonomic |
| Study design | RCT* or control or random* or "blind*" or "random* control*" |

### Articles selection

Abstracts from each database’ result list (PsycINFO and PubMed) were extracted and imported into EndNote X9, where duplicates were removed. Titles and abstracts were screened for inclusion criteria. Shortlisted studies, available as full text, were downloaded from the databases and further screened to ensure their relevance to the question and their adherence to the eligibility criteria discussed above. Screening of title/abstract and full text were performed by two independent reviewers (CF and AB).

### Data extraction and analysis

From full text, the following data was extracted: age of participants, conditions, settings (e.g., school, hospital), duration of intervention, stress biomarkers and main findings (significant effect of DAI at alpha .05). Descriptive analysis included outcomes of DAI on stress biomarkers versus no intervention, and other types of intervention, as well as effects on pre and post interventions. Effects of DAI in relation to participants’ stage of life (children, adolescent, adult, older adults) and settings were also discussed.

## Results

A total of 160 articles was obtained using the search strategy. Eleven duplicates were found and discarded. Forty-two studies were retained after reading titles and abstracts to screen for relevance. Most studies removed based on the selection criteria were summaries, protocols, or meta-analyses. A further 16 articles were removed after reading the full text as they did not meet all criteria (e.g., not including an animal-assisted intervention, not measuring stress). A total of 26 studies entered the systematic review. Figure 1 shows the different steps of the selection process. It should be noted that two of the included studies by Krause-Parello et al. utilize the exact same study population, most likely analyzing the same data.

**Figure 1.**
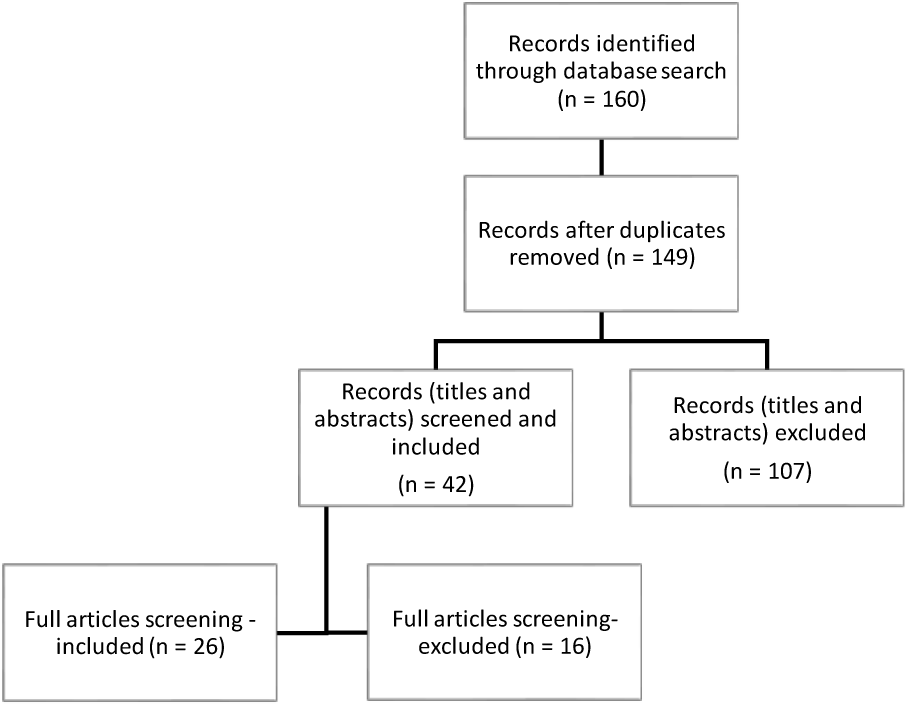
Flow Chart of Decisions Regarding Studies Included in the Systematic Review.

### Participants

A total of 1,829 participants were analyzed across studies. The participants either received a brief DAI or an alternative intervention (e.g. music therapy, art therapy, relaxation, or no intervention, or normal procedure in the setting). In two studies, participants received DAI and control intervention as part of the cross-over study design. Details about participants and conditions are shown in Table 1. 12 studies assessed explicitly the effect of AAI on children and teenagers who ranged from 2 to 19 years old. Two studies focused on older adults, leading to a maximum age of 86 years.

### Settings

The studies included were conducted in a wide range of settings. Fifteen were conducted in healthcare settings, five in laboratories, five in college or universities, and three at a trauma victim center.

### Stress Biomarkers

Different “wet” and “dry” biomarkers were reported to assess stress objectively in participants as shown in Fig. 2. Cortisol level, measured in saliva or blood, was the most frequently analyzed biomarker and was included in 16 out of the 26 studies. All but six studies used more than one biomarker. Other methods were more specific to the setting. For example, C-reactive protein, an inflammatory marker linked to psychological stress (Khandaker et al., 2016), was collected in hospitalized children who participated in Branson et al.’s (2017) study. Similarly, the study by Vagnoli et al. (2015) used plasma to measure stress related to venipuncture.

**Figure 2.**
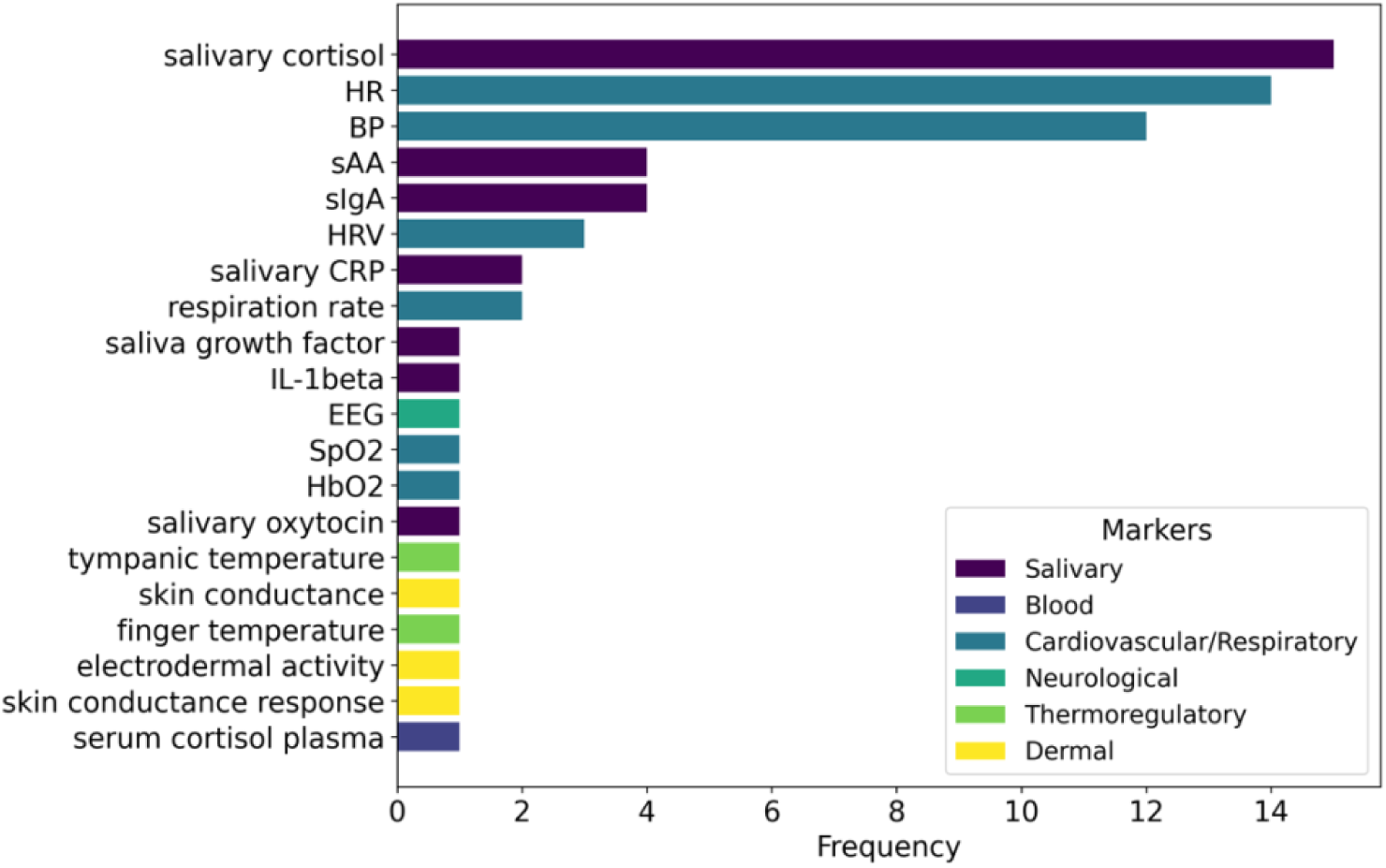
Frequency of Different Stress Biomarkers Used in Reviewed Studies.

### Investigating the Interventions

Most studies, 11 out of 13, reported a statistically significant (*p* < .05) effect of DAI on the measured changes in stress biomarkers. Over half (n = 6) the studies that measured cortisol levels reported a significant reduction in the DAI groups when compared to control groups. The findings were not specific to a population and effects were found across all settings. Similarly, four studies found an effect of DAI on heart rate, and results extended across populations and multiple settings. Two studies found a significant effect on immunoglobulin A levels. Interestingly, Gebhart et al. (2020) found that DAI significantly increased immunoglobulin A levels in students before exams, while Krause-Parello et al. (2018) found that children who received DAI during a forensic interview for allegations of child sexual abuse had significantly lower immunoglobulin A levels post interview than those who received a standard interview. Effects were also found with oxytocin, tympanic membrane temperature, but these were each found in single studies. The only study specific to adolescents found no statistically significant effect of DAI (Mueller et al., 2021). Only three studies included a group that received comparative therapeutic interventions, notably art or music therapy, or relaxation.

**Table 2.**
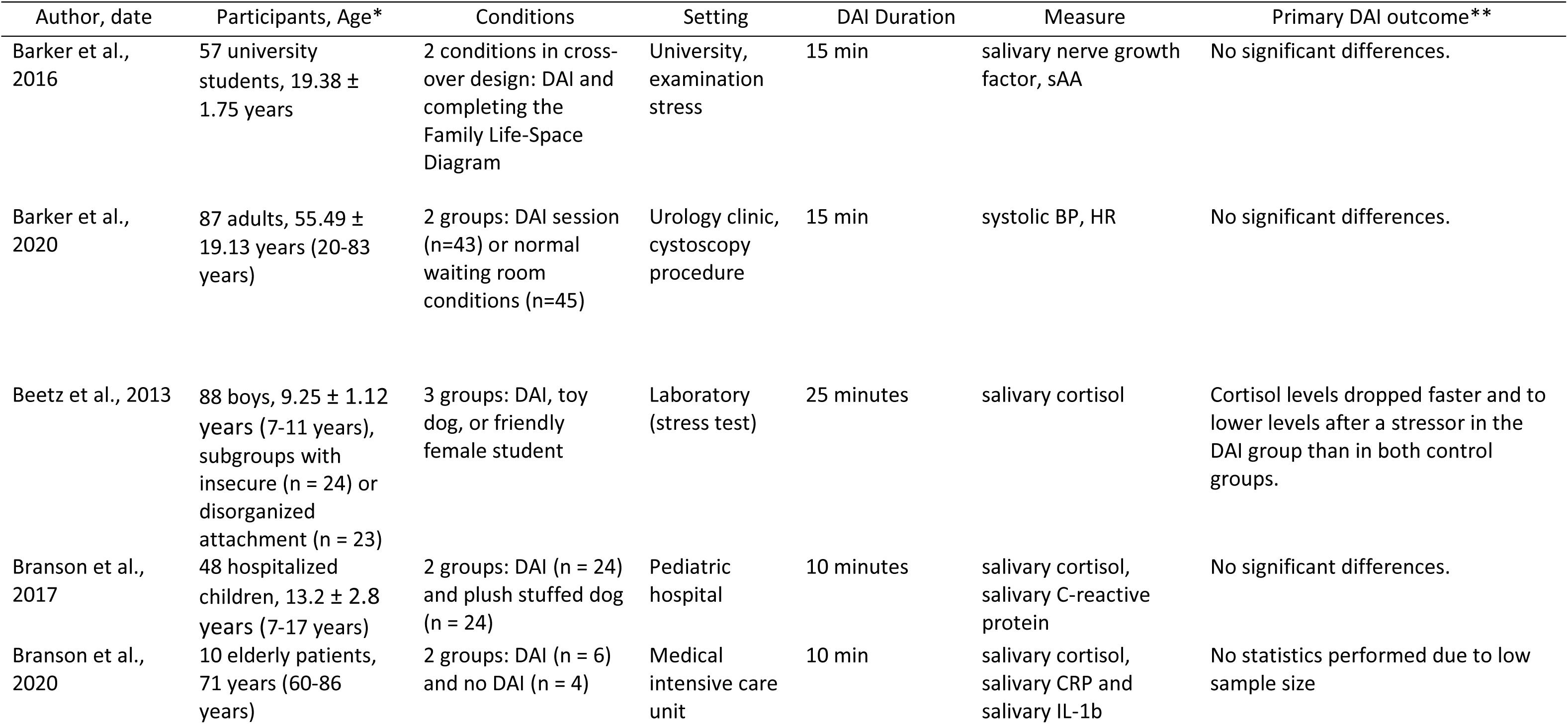
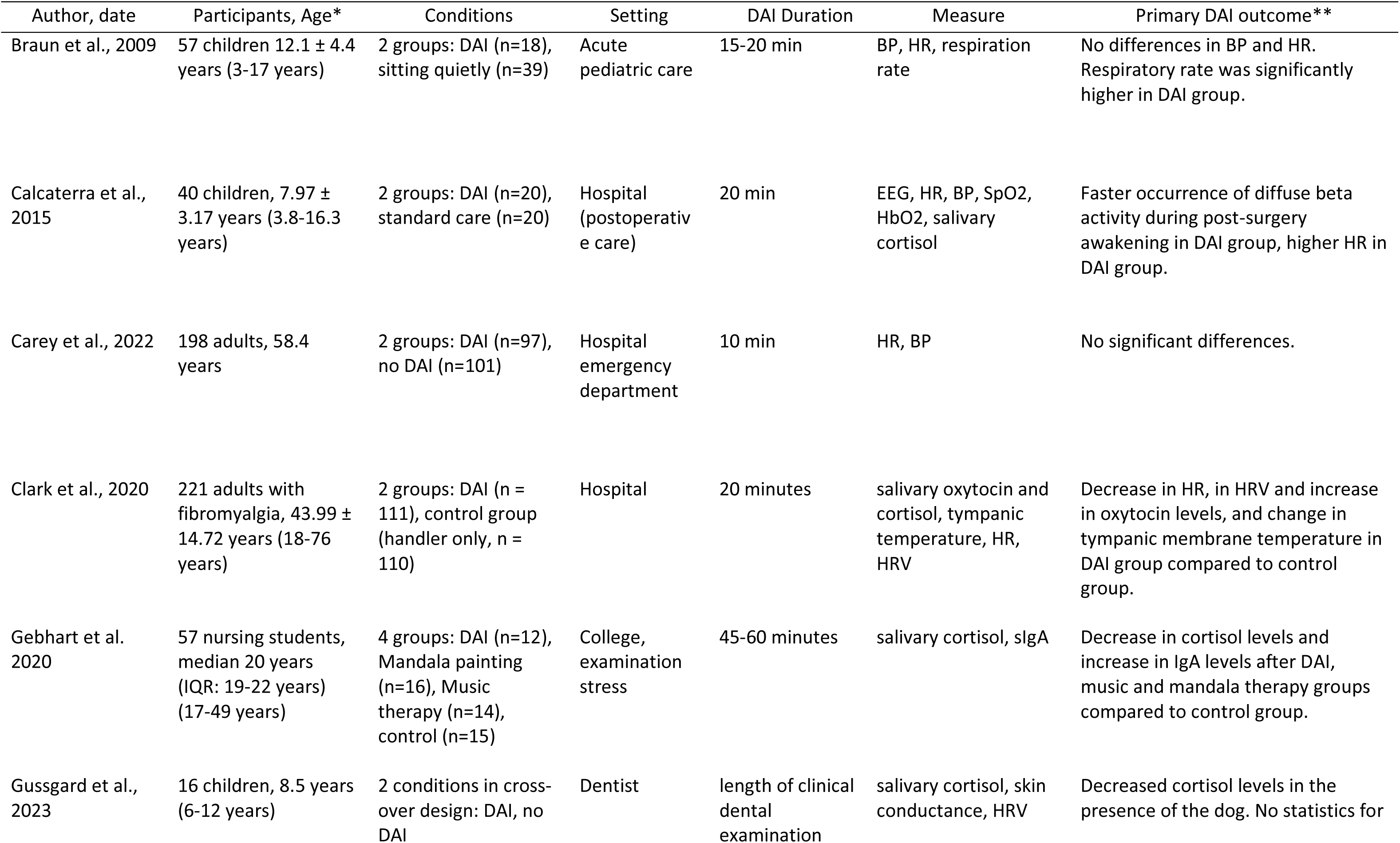
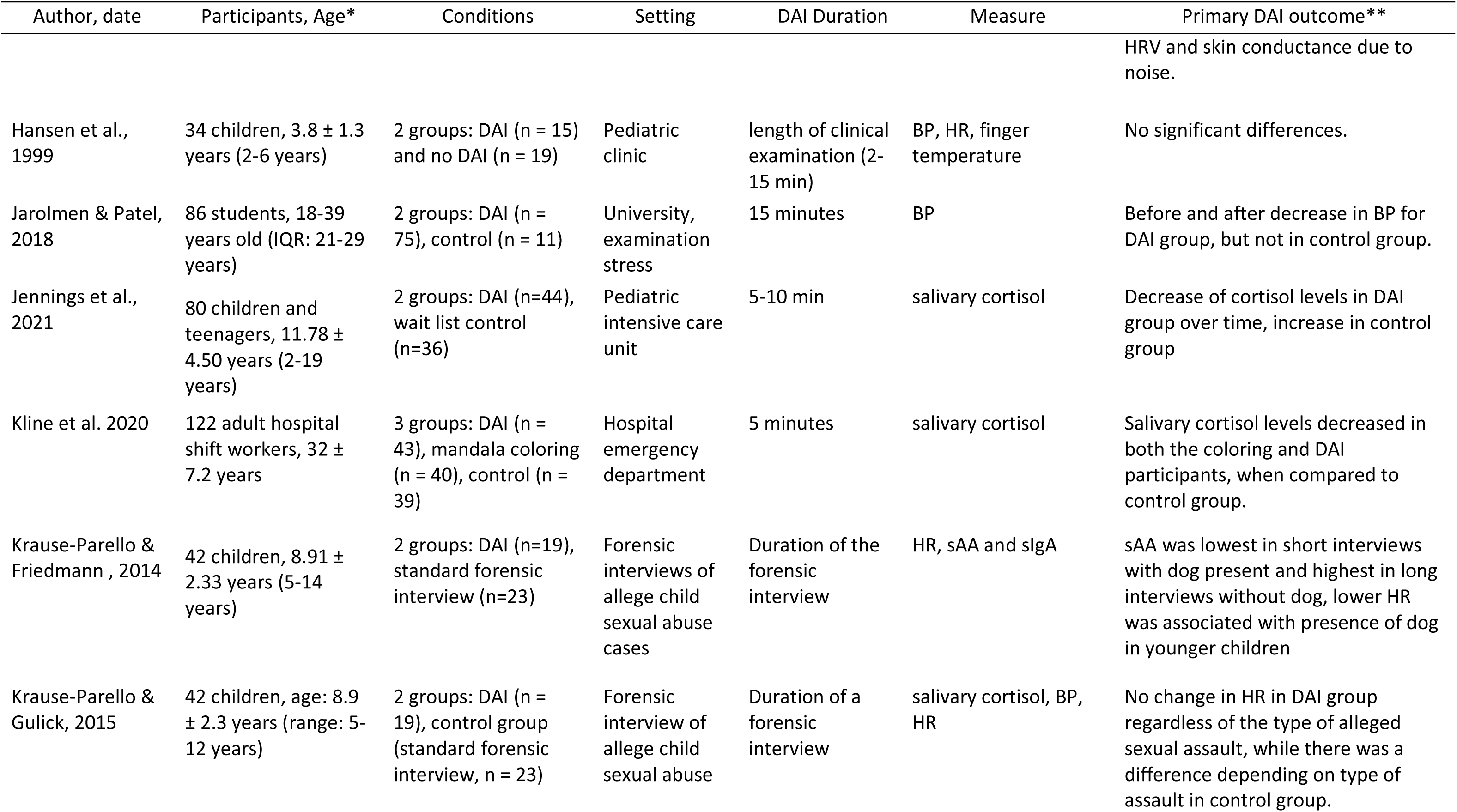
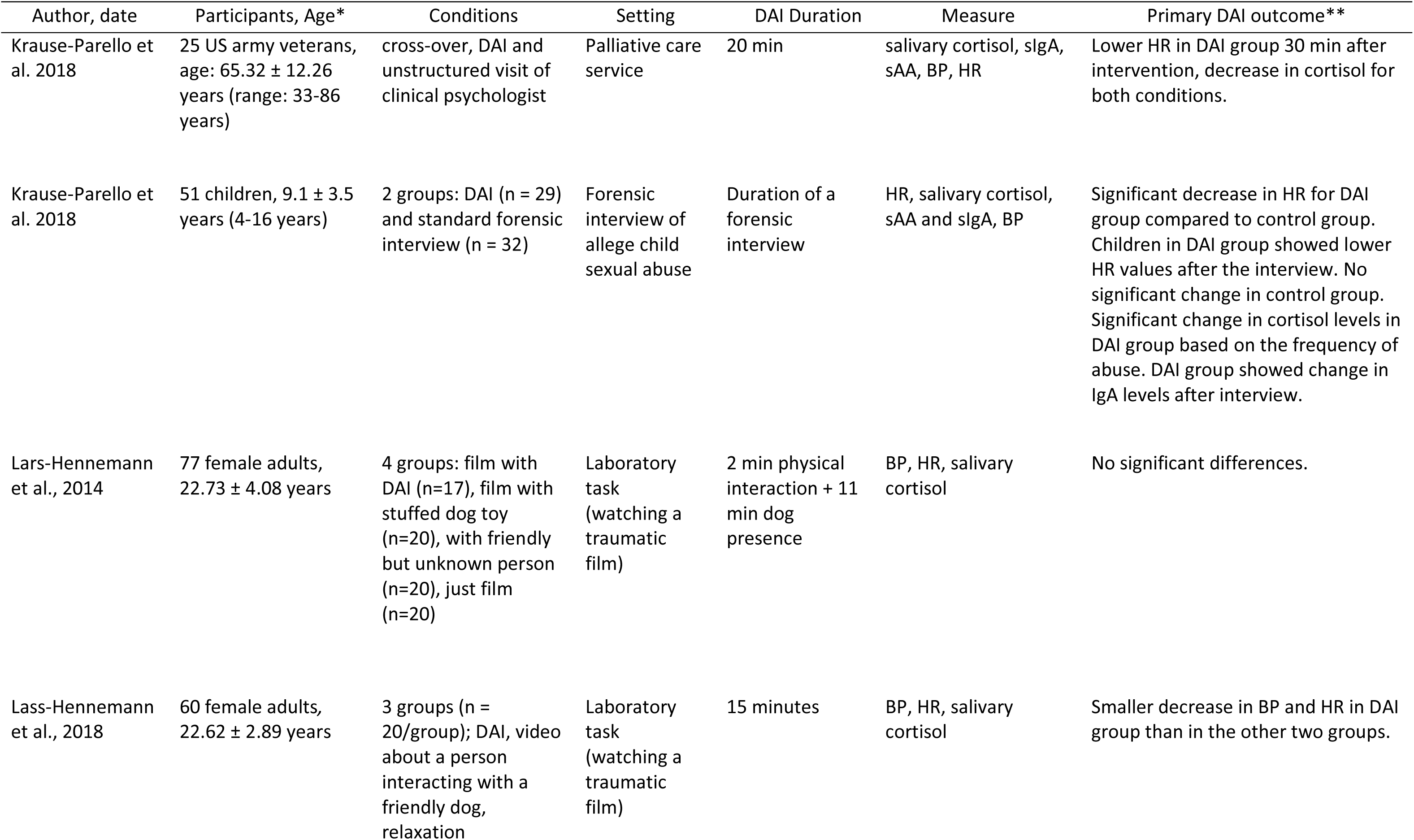
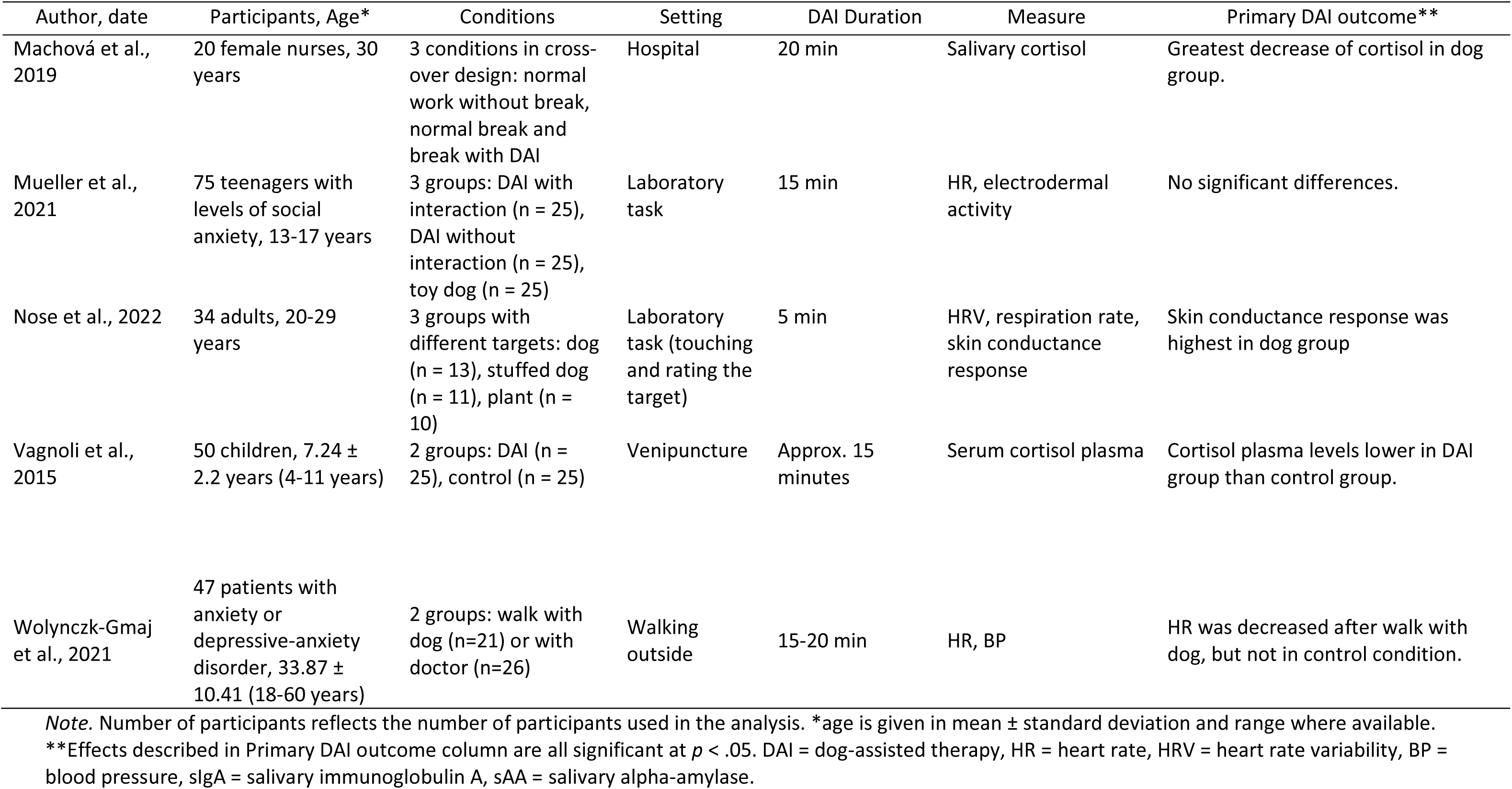
Summary of Studies Included in the Systematic Review.

## Discussion

The effectiveness of animal-assisted interactions has recently been under scrutiny, with leading researchers describing the field as biased (Fine et al., 2019). This systematic review narrowed in on high-quality empirical evidence of the changes in the physiological processes underpinning stress, and their use as objective measures of the therapeutic benefits of brief interventions with dogs, only. Evidence to date partially supports the notion that a brief therapeutic intervention with the assistance of a dog may influence the autonomic stress response given that the majority of studies found significant changes in stress biomarkers in groups receiving DAI compared to control groups.

### DAI and Cortisol Levels

Of the 20 stress biomarkers identified, changes in cortisol levels provided the most robust evidence. Prior literature demonstrated that when a person is exposed to certain stressors, the hypothalamic-pituitary-adrenal axis is activated, which causes blood cortisol levels to rise. After the stressor has passed, cortisol levels typically return to their baseline levels. For example, in a study investigating stress in younger adults, Cay et al. (2018) found that cortisol levels were nine times higher in a stressful situation than in a relaxing condition. Significant changes in cortisol levels were found in six of the studies in this review. Beetz et al. (2013), who investigated the effect of DAI on younger children with insecure or disorganized attachment style, found that cortisol levels dropped significantly faster and to lower levels after a stressor in the group with a dog-assisted intervention. They also found that the more time children pet the dog, the greater the drop in cortisol levels, suggesting a potential dose-dependent effect. Similar results using cortisol as a biomarker in studies of other types of stress reduction interventions have also been obtained, indicating that cortisol may be a reliable marker to assess the effect of brief therapeutic interventions on stress. For example, Hunter et al. (2019) investigated changes in cortisol levels following nature exposure. The authors found that nature exposure resulted in reduced cortisol levels over several hours following exposure, with the greatest rate of reduction observed between 20 and 30 minutes after exposure.

Nonetheless, several studies in this review reported no statistically significant effect of DAI on cortisol levels. While not included in the review, Crump and Derting (2015) suggested that pet ownership may affect results of DAI due to habituation to the benefits of interactions with animal. However, Branson et al., (2017) had considered pet ownership in their design and still did not observe a significant effect. An alternative explanation may stand in the type of stressor participants were exposed to prior to DAI. In the study by Branson, hospitalized children had baseline cortisol levels that were within the normal range expected for children aged 12–18 years (0.021–0.883 μg/dL; Salimetrics, 2016). As cortisol levels did not reduce significantly after the intervention, the authors suggested a possible ceiling effect. This aligns with other studies that found a significant change under a more stressful situation. For instance, Krause-Parello et al. (2019) found that cortisol decreased significantly (*p* < .05) in the DAI group as compared to the control group in military personnel following an aeromedical evacuation. Similarly, Vagnoli et al. (2015) found that cortisol levels were significantly lower in children undergoing venipuncture in the DAI than in the control group.

### The Use of HR and BP to Measure the Influence of DAI

Many studies employed BP and HR to objectively assess the effect of DAI on stress. These metrics offer several advantages. They are often cost-effective, easily accessible, and convenient in comparison to other measures requiring blood samples or saliva, which may require specialized material and more complex procedures. Nevertheless, our review highlights mixed results. Notably, even when significant differences were found, the direction was not always consistent. For instance, Krause-Parello et al. (2016) found a lower HR in DAI group 30 minutes after the intervention in a group of army veterans. In contrast, Calcaterra et al. (2015) found an increase in HR in the DAI group during the intervention in children. This difference in results suggests that the timing of collection may play a key role in the interpretation of these metrics. When measured during the intervention, it is possible that arousal from interacting with an animal may not indicate elevated distress, but rather excitement which would have a similar effect on HR. The timing of the intervention, before or after the stressful event, may also influence these results as one may measure how DAI may assist in reducing distress during a stressful event, while the other may measure the effect DAI on recovery. Variation in participants may also be contributing to the heterogeneity of the results (e.g, children’s reaction to dogs in comparison with adults). Standardization in methodologies when collecting these would help clarify the relationship between DAI and physiological stress responses.

### DAI Compared to Other Therapeutic Interventions

The results of the review indicate that there is insufficient evidence to confidently evaluate whether DAI is more effective than other therapeutic interventions in reducing physiological stress. In the three studies that had included other therapeutic modalities, the conditions and biomarkers used were too dissimilar to draw conclusions from the mixed results produced. For instance, Lass-Hennemann et al. (2018) found that a smaller decrease in heart rate and blood pressure with DAI than in the relaxation condition. While the decrease in cardiovascular measures can be interpreted as a reduction in stress during a relaxation condition, it could also indicate pleasant arousal in the DAI condition, therefore failing to measure the same construct in each condition. Results from Kline et al. (2020) indicated that salivary cortisol levels decreased significantly in both the art and dog assisted therapies. However, only the DAI condition was associated with a reduction in reported stress (as measured by a visual analogue scale of stress) by the end of the shift. As discussed earlier, the perceived stress effect of DAI has frequently been reported as being a strength of animal-assisted therapy. This was supported by a meta-analysis that found significant changes in self-reported measures of stress before and after pet therapy (Ein et al., 2018).

### DAI Applied to Different Groups and Contexts

Due to the heterogeneity of the studies, the present review failed to reach a conclusion regarding the effect of DAI on stress biomarkers within a specific population or settings. Still, preliminary evidence suggests that DAI may be suitable across the lifespan due to significant effects being found at least once in all age groups. While the study specific to adolescent participants found no effect, the age range partially overlapped with the ages of children included in studies that measured a change (e.g., Krause-Parello et al., 2018). Similarly, effects of DAI on biomarkers were found across all settings, indicating that DAI may offer therapeutic benefits in several contexts including health, education, and trauma. Our conclusion is in line with another systematic review that focused on trauma, which suggested that while therapy with the assistance of animals may be promising, more rigid research protocols are needed to truly measure clinical efficacy (O’Haire et al., 2015).

### Limitations and Strengths of the Current Review

This review has several limitations. Most importantly, the variety of biomarkers used in the studies, as well as the different protocol and AAI terminology made it difficult to assess the magnitude of the effect of AAI as an intervention for psychological stress. Lack of methodological rigor and transparency of several studies limited our ability to reach firm conclusions. To address this issue, future systematic reviews should assess the quality of RTCs with a valid and reliable scale (Olivo et al., 2008), and only include those of high standards in their analysis.

Nonetheless, the current review narrowed in on specific physiological parameters that lend strength to the argument of DAI as an effective therapy to reduce biological stress. As a result, there is sound evidence to continue further investigation into the use of biomarkers such as cortisol and heart rate to measure the effectiveness of DAI in situations of acute stress.

### Clinical Implications and Future Directions

Stress reduction interventions with the assistance of dogs are already in place in many settings and populations. DAI offers a highly appealing therapeutic option which, as discussed earlier, is thought to be rooted in the human-animal bond. The current review provides preliminary evidence of the effect of DAIs on the autonomic stress response, thereby supporting its efficacy as a brief intervention, particularly in acute stress situations. Most importantly, the review identified possible reliable biomarkers, cortisol levels, BP and HR which give a clear direction for future studies. More data is required to confirm their validity and associations with self-reported stress measures. Furthermore, this review calls for this combined method to be applied in gold-standard RCTs to compare repeated long-term DAI with other long-term stress reduction interventions. Considering the disastrous health consequences of stress, incorporating dogs into other stress therapy may provide a simple and appealing way to encourage people to access clinical psychological services.

## Data Availability

All data produced in the present work are contained in the manuscript

## References

American Psychological Association. (n.d.). Stress. In APA dictionary of psychology. Retrieved October 18, 2020, from https://dictionary.apa.org/stress

Bagheri, M., & Power, S. D. (2020). EEG-based detection of mental workload level and stress: the effect of variation in each state on classification of the other. Journal of Neural Engineering, 17(5), 056015. 10.1088/1741-2552/abbc27

Barker, S. B., Barker, R. T., McCain, N. L., & Schubert, C. M. (2016). A randomized cross-over exploratory study of the effect of visiting therapy dogs on college student stress before final exams [Empirical Study; Quantitative Study]. Anthrozoos, 29(1), 35–46. 10.1080/08927936.2015.1069988

Barker, S., Krzastek, S., Vokes, R., Schubert, C., Cooley, L. F., & Hampton, L. J. (2020). Examining the effect of an animal-assisted intervention on patient distress in outpatient cystoscopy [Empirical Study; Quantitative Study]. Human-Animal Interaction Bulletin, 8(1), 23–37.

Beck, A. M. (2014). The biology of the human–animal bond. Animal Frontiers, 4(3), 32–36. 10.2527/af.2014-0019

Beetz, A., Julius, H., Turner, D., & Kotrschal, K. (2012). Effects of social support by a dog on stress modulation in male children with insecure attachment [Empirical Study; Quantitative Study]. Frontiers in Psychology Vol 3 2012, ArtID 352, 3101:. 10.3389/fpsyg.2012.00352

Bowlby, J. (1958). The nature of the child’s tie to his mother. International Journal of Psychoanalysis, 39, 350–371.

Branson, S., Boss, L., Hamlin, S., & Padhye, N. S. (2020). Animal-Assisted Activity in Critically Ill Older Adults: A Randomized Pilot and Feasibility Trial. Biol Res Nurs, 22(3), 412–417. 10.1177/1099800420920719

Branson, S. M., Boss, L., Padhye, N. S., Trötscher, T., & Ward, A. (2017). Effects of Animal-assisted Activities on Biobehavioral Stress Responses in Hospitalized Children: A Randomized Controlled Study. J Pediatr Nurs, 36, 84–91. 10.1016/j.pedn.2017.05.006

Braun, C., Stangler, T., Narveson, J., & Pettingell, S. (2009). Animal-assisted therapy as a pain relief intervention for children. Complement Ther Clin Pract, 15(2), 105–109. 10.1016/j.ctcp.2009.02.008

Cay, M., Ucar, C., Senol, D., Cevirgen, F., Ozbag, D., Altay, Z., & Yildiz, S. (2018). Effect of increase in cortisol level due to stress in healthy young individuals on dynamic and static balance scores. North Clin Istanb, 5(4), 295–301. 10.14744/nci.2017.42103

Chambers, J., Quinlan, M. B., Evans, A., & Quinlan, R. J. (2020). Dog-Human Coevolution: Cross-Cultural Analysis of Multiple Hypotheses. Journal of Ethnobiology, 40, 414–433.

Chandler, C. K. (2012). Animal assisted therapy in counseling (2nd ed.). Routledge.

Clark, S., Martin, F., McGowan, R. T. S., Smidt, J., Anderson, R., Wang, L., Turpin, T., Langenfeld-McCoy, N., Bauer, B., & Mohabbat, A. B. (2020). The Impact of a 20-Minute Animal-Assisted Activity Session on the Physiological and Emotional States in Patients With Fibromyalgia. Mayo Clin Proc, 95(11), 2442–2461. 10.1016/j.mayocp.2020.04.037

Calcaterra, V., Veggiotti, P., Palestrini, C., De Giorgis, V., Raschetti, R., Tumminelli, M., Mencherini, S., Papotti, F., Klersy, C., Albertini, R., Ostuni, S., & Pelizzo, G. (2015). Post-operative benefits of animal-assisted therapy in pediatric surgery: a randomised study. PLoS One, 10(6), e0125813. 10.1371/journal.pone.0125813

Carey, B., Dell, C. A., Stempien, J., Tupper, S., Rohr, B., Carr, E., Cruz, M., Acoose, S., Butt, P., Broberg, L., Collard, L., Fele-Slaferek, L., Fornssler, C., Goodridge, D., Gunderson, J., McKenzie, H., Rubin, J., Shand, J., Smith, J., … Meier, S. (2022). Outcomes of a controlled trial with visiting therapy dog teams on pain in adults in an emergency department. PLoS One, 17(3), e0262599. 10.1371/journal.pone.0262599

Crump, C., & Derting, T. L. (2015). Effects of pet therapy on the psychological and physiological stress levels of first-year female undergraduates [Empirical Study; Quantitative Study]. North American Journal of Psychology, 17(3), 575–590.

Cullen, F. T., & Wilcox, P. (2010). Encyclopedia of Criminological Theory. SAGE Publications.

Ein, N., Li, L., & Vickers, K. (2018). The effect of pet therapy on the physiological and subjective stress response: A meta-analysis [Meta Analysis]. Stress and Health: Journal of the International Society for the Investigation of Stress, 34(4), 477–489. 10.1002/smi.2812

Fine, A. H., Beck, A. M., & Ng, Z. (2019). The State of Animal-Assisted Interventions: Addressing the Contemporary Issues that will Shape the Future. International journal of environmental research and public health, 16(20), 3997. 10.3390/ijerph16203997

Friedmann, E., Katcher, A. H., Lynch, J. J., & Thomas, S. A. (1980). Animal companions and one-year survival of patients after discharge from a coronary care unit. Public health reports (Washington, D.C. : 1974), 95(4), 307–312. https://pubmed.ncbi.nlm.nih.gov/6999524

Gebhart, V., Buchberger, W., Klotz, I., Neururer, S., Rungg, C., Tucek, G., Zenzmaier, C., & Perkhofer, S. (2020). Distraction-focused interventions on examination stress in nursing students: Effects on psychological stress and biomarker levels. A randomized controlled trial [Empirical Study; Quantitative Study]. International Journal of Nursing Practice Vol 26(1), 2020, ArtID e12788, 26(1). 10.1111/ijn.12788

Gordon, A. M., & Mendes, W. B. (2021). A large-scale study of stress, emotions, and blood pressure in daily life using a digital platform. Proceedings of the National Academy of Sciences, 118(31), e2105573118. 10.1073/pnas.2105573118

Grajfoner, D., Harte, E., Potter, L. M., & McGuigan, N. (2017). The Effect of Dog-Assisted Intervention on Student Well-Being, Mood, and Anxiety. International journal of environmental research and public health, 14(5), 483. 10.3390/ijerph14050483

Gussgard, A. M., Carlstedt, K., & Meirik, M. (2023). Intraoral clinical examinations of pediatric patients with anticipatory anxiety and situational fear facilitated by therapy dog assistance: A pilot RCT. Clin Exp Dent Res, 9(1), 122–133. 10.1002/cre2.679

Hek, K., Direk, N., Newson, R. S., Hofman, A., Hoogendijk, W. J. G., Mulder, C. L., & Tiemeier, H. (2013). Anxiety disorders and salivary cortisol levels in older adults: a population-based study. Psychoneuroendocrinology, 38(2), 300–305. 10.1016/j.psyneuen.2012.06.006

Hansen, K. M., Messinger, C. J., Baun, M. M., & Megel, M. (1999). Companion animals alleviating distress in children [Empirical Study]. Anthrozoos, 12(3), 142–148. 10.2752/089279399787000264

Horowitz, A. (2009). Disambiguating the “guilty look”: salient prompts to a familiar dog behaviour. Behav Processes, 81(3), 447–452. 10.1016/j.beproc.2009.03.014

Hunter, M. R., Gillespie, B. W., & Chen, S. Y.-P. (2019). Urban Nature Experiences Reduce Stress in the Context of Daily Life Based on Salivary Biomarkers [Original Research]. Frontiers in Psychology, 10(722). 10.3389/fpsyg.2019.00722

International Association of Human-Animal Interaction Organizations. (2018). The IAHAIO definitions for animal assisted intervention and guidelines for wellness of animals involved in AAI. https://iahaio.org/wp/wp-content/uploads/2021/01/iahaio-white-paper-2018-english.pdf

Jarolmen, J., & Patel, G. (2018). The effects of animal-assisted activities on college students before and after a final exam [Empirical Study; Quantitative Study]. Journal of Creativity in Mental Health, 13(3), 264–274. 10.1080/15401383.2018.1425941

Jennings, M. L., Granger, D. A., Bryce, C. I., Twitchell, D., Yeakel, K., & Teaford, P. A. (2021). Effect of animal assisted interactions on activity and stress response in children in acute care settings. Compr Psychoneuroendocrinol, 8, 100076. 10.1016/j.cpnec.2021.100076

Khan, Q. U. (2020). Relationship of Salivary Cortisol Level With Severe Depression and Family History. Cureus, 12(11), e11548–e11548. 10.7759/cureus.11548

Khandaker, G. M., Zammit, S., Lewis, G., & Jones, P. B. (2016). Association between serum C-reactive protein and DSM-IV generalized anxiety disorder in adolescence: Findings from the ALSPAC cohort. Neurobiology of Stress, 4, 55–61. 10.1016/j.ynstr.2016.02.003

Kline, J. A., VanRyzin, K., Davis, J. C., Parra, J. A., Todd, M. L., Shaw, L. L., Haggard, B. R., Fisher, M. A., Pettit, K. L., & Beck, A. M. (2020, Apr). Randomized Trial of Therapy Dogs Versus Deliberative Coloring (Art Therapy) to Reduce Stress in Emergency Medicine Providers. Acad Emerg Med, 27(4), 266–275. 10.1111/acem.13939

Krause-Parello, C. A., & Friedmann, E. (2014). The effects of an animal-assisted intervention on salivary alpha-amylase, salivary immunoglobulin a, and heart rate during forensic interviews in child sexual abuse cases [Empirical Study; Interview; Quantitative Study]. Anthrozoos, 27(4), 581–590. 10.2752/089279314X14072268688005

Krause-Parello, C. A., & Gulick, E. E. (2015). Forensic interviews for child sexual abuse allegations: An investigation into the effects of animal-assisted intervention on stress biomarkers [Empirical Study; Quantitative Study]. Journal of Child Sexual Abuse: Research, Treatment, & Program Innovations for Victims, Survivors, & Offenders, 24(8), 873–886. 10.1080/10538712.2015.1088916

Krause-Parello, C. A., Levy, C., Holman, E., & Kolassa, J. E. (2018). Effects of VA Facility Dog on Hospitalized Veterans Seen by a Palliative Care Psychologist: An Innovative Approach to Impacting Stress Indicators. Am J Hosp Palliat Care, 35(1), 5–14. 10.1177/1049909116675571

Krause-Parello, C. A., Thames, M., Ray, C. M., & Kolassa, J. (2018). Examining the effects of a service-trained facility dog on stress in children undergoing forensic interview for allegations of child sexual abuse [Empirical Study; Quantitative Study]. Journal of Child Sexual Abuse: Research, Treatment, & Program Innovations for Victims, Survivors, & Offenders, 27(3), 305–320. 10.1080/10538712.2018.1443303

Lass-Hennemann, J., Peyk, P., Streb, M., Holz, E., & Michael, T. (2014). Presence of a dog reduces subjective but not physiological stress responses to an analog trauma. Front Psychol, 5, 1010. 10.3389/fpsyg.2014.01010

Lass-Hennemann, J., Schafer, S. K., Romer, S., Holz, E., Streb, M., & Michael, T. (2018). Therapy dogs as a crisis intervention after traumatic events?-An experimental study [Empirical Study; Quantitative Study]. Frontiers in Psychology Vol 9 2018, ArtID 1627, 9. 10.3389/fpsyg.2018.01627

Machová, K., Součková, M., Procházková, R., Vaníčková, Z., & Mezian, K. (2019). Canine-Assisted Therapy Improves Well-Being in Nurses. Int J Environ Res Public Health, 16(19). 10.3390/ijerph16193670

Mariotti, A. (2015). The effects of chronic stress on health: new insights into the molecular mechanisms of brain-body communication. Future science OA, 1(3), FSO23–FSO23. 10.4155/fso.15.21

McCullough, A., Ruehrdanz, A., Jenkins, M. A., Gilmer, M. J., Olson, J., Pawar, A., Holley, L., Sierra-Rivera, S., Linder, D. E., Pichette, D., Grossman, N. J., Hellman, C., Guerin, N. A., & O’Haire, M. E. (2018). Measuring the effects of an animal-assisted intervention for pediatric oncology patients and their parents: A multisite randomized controlled trial [Empirical Study; Quantitative Study; Treatment Outcome]. Journal of Pediatric Oncology Nursing, 35(3), 159–177. 10.1177/1043454217748586

Mueller, M. K., Anderson, E. C., King, E. K., & Urry, H. L. (2021). Null effects of therapy dog interaction on adolescent anxiety during a laboratory-based social evaluative stressor. Anxiety, Stress & Coping: An International Journal, No Pagination Specified. 10.1080/10615806.2021.1892084

Nose, I., Masamoto, K., Tsuchida, A., Hayashi, M., Irimajiri, M., & Kakinuma, M. (2022). The effect of interaction with a dog on heart rate variability based on Lorenz plot analysis [Empirical Study; Quantitative Study]. Human-Animal Interaction Bulletin, 10(1), 84–99.

O’Haire, M. E., Guérin, N. A., & Kirkham, A. C. (2015). Animal-Assisted Intervention for trauma: a systematic literature review. Front Psychol, 6, 1121. 10.3389/fpsyg.2015.01121

Odendaal, J. S. J. (2001, Mar). A physiological basis for animal-facilitated psychotherapy [Dissertation Empirical Study]. Dissertation Abstracts International: Section B: The Sciences and Engineering, 61(9-B), 4999.

Olivo, S. A., Macedo, L. G., Gadotti, I. C., Fuentes, J., Stanton, T., & Magee, D. J. (2008). Scales to Assess the Quality of Randomized Controlled Trials: A Systematic Review. Physical Therapy, 88(2), 156–175. 10.2522/ptj.20070147

Page, M. J., McKenzie, J. E., Bossuyt, P. M., Boutron, I., Hoffmann, T. C., Mulrow, C. D., Shamseer, L., Tetzlaff, J. M., Akl, E. A., Brennan, S. E., Chou, R., Glanville, J., Grimshaw, J. M., Hróbjartsson, A., Lalu, M. M., Li, T., Loder, E. W., Mayo-Wilson, E., McDonald, S., McGuinness, L. A., Stewart, L. A., Thomas, J., Tricco, A. C., Welch, V. A., Whiting, P., & Moher, D. (2021). The PRISMA 2020 statement: an updated guideline for reporting systematic reviews. Bmj, 372, n71. 10.1136/bmj.n71

Satti, F. A., Hussain, M., Hussain, J., Kim, T. S., Lee, S., & Chung, T. (2021, 4-6 Jan. 2021). User Stress Modeling through Galvanic Skin Response. 2021 15th International Conference on Ubiquitous Information Management and Communication (IMCOM).

Taelman, J., Vandeput, S., Spaepen, A., & Van Huffel, S. (2009). Influence of Mental Stress on Heart Rate and Heart Rate Variability. 4th European Conference of the International Federation for Medical and Biological Engineering, Berlin, Heidelberg.

Vagnoli, L., Caprilli, S., Vernucci, C., Zagni, S., Mugnai, F., & Messeri, A. (2015). Can presence of a dog reduce pain and distress in children during venipuncture? [Empirical Study; Quantitative Study]. Pain Management Nursing, 16(2), 89–95. 10.1016/j.pmn.2014.04.004

Wilson, E. O. (1986). Biophilia. Harvard University Press. 10.4159/9780674045231

Wołyńczyk-Gmaj, D., Ziółkowska, A., Rogala, P., Ścigała, D., Bryła, L., Gmaj, B., & Wojnar, M. (2021). Can Dog-Assisted Intervention Decrease Anxiety Level and Autonomic Agitation in Patients with Anxiety Disorders? J Clin Med, 10(21). 10.3390/jcm10215171

